# Household Transmission and Clinical Features of SARS-CoV-2 Infections by Age in 2 US Communities

**DOI:** 10.1101/2021.08.16.21262121

**Authors:** Huong Q. McLean, Carlos G. Grijalva, Kayla E. Hanson, Yuwei G. Zhu, Jessica E. Deyoe, Jennifer K. Meece, Natasha B. Halasa, James D. Chappell, Alexandra Mellis, Carrie Reed, Edward A. Belongia, H. Keipp Talbot, Melissa A. Rolfes

**Affiliations:** Marshfield Clinic Research Institute, Marshfield, Wisconsin; Vanderbilt University Medical Center, Nashville, Tennessee; Centers for Disease Control and Prevention, Atlanta, Georgia

## Abstract

**OBJECTIVES:** Examine age differences in SARS-CoV-2 transmission risk from primary cases and infection risk among household contacts, and symptoms among those with SARS-CoV-2 infection.

**METHODS:** People with SARS-CoV-2 infection in Nashville, Tennessee and central and western Wisconsin and their household contacts were followed daily for 14 days to ascertain symptoms and secondary transmission events. Households were enrolled between April 2020 and April 2021. Secondary infection risks (SIR) by age of the primary case and contacts were estimated using generalized estimating equations.

**RESULTS:** The 226 primary cases were followed by 198 (49%) secondary SARS-CoV-2 infections among 404 household contacts. Age group-specific SIR among contacts ranged from 36% to 53%, with no differences by age. SIR was lower from primary cases aged 12-17 years than from primary cases 18-49 years (risk ratio [RR] 0.42; 95% confidence interval [CI] 0.19-0.91). SIR was 55% and 45%, respectively, among primary case-contact pairs in the same versus different age group (RR 1.47; 95% CI 0.98-2.22). SIR was highest among primary case-contacts pairs aged ≥65 years (76%) and 5-11 years (69%). Among secondary SARS-CoV-2 infections, 19% were asymptomatic; there was no difference in the frequency of asymptomatic infections by age group.

**CONCLUSIONS:** Both children and adults can transmit and are susceptible to SARS-CoV-2 infection. SIR did not vary by age, but further research is needed to understand age-related differences in probability of transmission from primary cases by age.

## BACKGROUND

While some studies suggest children (age <18 years) are less susceptible to SARS-CoV-2 infection than adults,^1-4^ other studies of household or other close contacts have found similar secondary infection rates among children and adults.^5,6^ Variation in mixing patterns, and likelihood of exposure and detection (contact tracing and testing practices) may contribute to reported differences in children versus adults.^2,7^ Most prior reports on SARS-CoV-2 infection in children utilized surveillance reports or contact tracing data, and were conducted early in the course of the pandemic. During that time, children were largely protected from community exposures due to non-pharmaceutical interventions, including closure of businesses and schools. Furthermore, many of those early studies were conducted in Asia, where prevention efforts and age-related interactions likely differed from those in the United States (US).^1-3^

Children tend to have less severe illness than adults,^8-11^ but the spectrum of illness and SARS-CoV-2 transmission risk have not been fully characterized among children in US households. Furthermore, little is known about the risk in the youngest age groups. Greater understanding of age-related differences in susceptibility, transmission risk, and illness characteristics, particularly in children, is needed to guide public health recommendations on prevention of transmission and inform plans to resume in-person school attendance.

We previously reported on SARS-CoV-2 transmission from a prospective study of US households.^12^ We extended those results and examined age differences in SARS-CoV-2 transmission risk from primary cases and infection risk among household contacts. We also assessed age-specific differences in symptoms and illness duration among secondary cases.

## METHODS

### Design, setting, and participants

This analysis used systematically and frequently collected data from a prospective case-ascertained household SARS-CoV-2 transmission study conducted in Tennessee and Wisconsin.^12,13^ Persons with laboratory-confirmed SARS-CoV-2 infection (index cases) were identified from SARS-CoV-2 clinical real-time reverse transcription polymerase chain reaction (rRT-PCR) tests conducted at Vanderbilt University Medical Center (VUMC; Nashville, Tennessee) and Marshfield Clinic Health System (MCHS; Marshfield, Wisconsin). VUMC is a large healthcare provider system serving patients from Tennessee and the Mid-South US. For this study, VUMC enrolled patients presenting to the network of walk-in-clinics that operate within Davidson County and surrounding areas. MCHS is a large community-based, multispecialty healthcare system serving predominantly rural populations in central, northern, and western Wisconsin. For this study, participants were recruited from MCHS locations in central and western Wisconsin. All or most schools in the study area were closed to in-person attendance in Spring 2020. In Fall 2020, some schools (public and private in Wisconsin and private in Tennessee) were open for full-time in-person attendance or had hybrid modalities (combination of in-person/remote attendance). SARS-CoV-2 testing capacity at both sites varied throughout the study period. Testing was limited in Spring 2020, increased by Summer 2020, with return times for results taking longer during periods with high level of community transmission. By Fall 2020, SARS-CoV-2 testing services were readily available with rapid return of results (same or next day in most cases).^14^

Index cases and their household contacts were followed daily for 14 days to ascertain symptoms and secondary transmission events. Household members were eligible if the index case had symptom onset <7 days before enrollment and there was ≥1 other household member without symptoms at the time of the index case’s illness onset. The primary case was the person with laboratory-confirmed SARS-CoV-2 infection in the household with the earliest illness onset date (or date of positive SARS-CoV-2 sample, if asymptomatic). This analysis included households enrolled between April 21, 2020 and April 30, 2021.

### Data and sample collection

Data were primarily collected through self- or parent-administered paper (Wisconsin) or web-based surveys (Tennessee); some data were obtained through interviews with participants. At enrollment, the survey assessed demographic and household characteristics, pre-existing medical conditions, occupational risk (e.g., employed in healthcare setting or customer service), symptoms prior to enrollment, and type and frequency of interactions with other household members. Each day during the 14-day follow-up period, participants were asked about current symptoms and provided a (self- or parent-collected) respiratory (anterior nasal) and/or saliva sample for SARS-CoV-2 rRT-PCR testing regardless of symptoms. Symptoms assessed included: constitutional symptoms (chills, fatigue or feeling run down, fever or feverishness, muscle or body aches), upper respiratory symptoms (nasal congestion, runny nose, sore throat), lower respiratory symptoms (chest tightness or pain, cough, trouble breathing or shortness of breath, wheezing), neurologic symptoms (headache, loss of taste or smell), and gastrointestinal symptoms (abdominal pain, diarrhea, vomiting). All participants were asked about all symptoms except gastrointestinal, which was only included in Wisconsin.

### Laboratory

Respiratory and saliva samples were tested using Center for Disease Control and Prevention (CDC), Quidel Lyra, or ThermoFisher Taq Path SARS-CoV-2 rRT-PCR assays and protocols at MCHS’ Research Institute or VUMC.^15-17^

### Analysis

Participants were grouped by age reflecting potential exposure risk and behavioral characteristics: preschool-aged (0-4 years), primary school-aged (5-11 years), secondary school-aged (12-17 years), young adults (18-49 years), middle-aged adults (50-64 years), and older adults (≥65 years).

Differences among age groups were assessed using Chi-square test or Kruskal-Wallis test, where appropriate. Secondary SARS-CoV-2 infections were defined as household contacts with ≥1 rRT-PCR positive sample (respiratory or saliva) with illness onset or first positive sample date within 14 days after the illness onset (or date of first positive sample, if asymptomatic) in the primary case. Secondary infection risks (SIR) were estimated using generalized estimating equations (GEE, log-binomial model), accounting for household clustering. We estimated SIR 1) by age of the primary case to assess transmission risk (the probability of transmission from the primary case to contacts), 2) by age of the contacts to assess infection risk (the probability of infection among contacts), 3) by age of the primary case and age of the contacts, and 4) by whether the primary case and contact were in the same or different age group. Risk ratios (RR) and 95% confidence intervals (CI) were used to compare SIR in each primary and contact age group versus age 18-49 years (referent) and same versus different age groups. Households with ≥1 co-primary cases (household members positive for SARS-CoV-2 that had illness onset within 2 days after illness onset in the primary case) were excluded.

We assessed frequency, sequence, and duration of symptoms by age group among participants with SARS-CoV-2 infection whose illness onset or first positive sample (if asymptomatic) occurred after study enrollment. Associations between age group and presence of specific categories of symptoms were assessed using logistic regression models. Odds ratios (OR) and 95% CI were used to compare odds of symptoms in each age group versus age 18-49 years (referent). Neurologic symptoms were not assessed in children aged 0-4 years, as these symptoms are difficult to ascertain in young children. Analysis of symptom duration and interval (in days) between symptom onset and first positive sample was restricted to symptomatic infections. Analyses were performed using SAS 9.4 (SAS Institute Inc., Cary, NC). The study protocol was approved by Institutional Review Boards at VUMC and MCHS. CDC determined this activity was conducted consistent with applicable federal law and CDC policy (see 45 C.F.R. part 46; 21 C.F.R. part 56).

## RESULTS

### Participant characteristics

From April 2020 through April 2021, 302 index patients and their 577 household members were enrolled. The index patient was the primary case for 96% of households. The primary analysis included 226 of 302 (75%) households with 404 contacts. Reasons for exclusion included: receipt of COVID-19 vaccine before enrollment (n=28) or contacts of a vaccinated primary case (n=4), illness onset >10 days before enrollment (n=15) as infections may have become undetectable, illness onset before the primary case without laboratory-confirmed SARS-CoV-2 infection (n=16), <6 follow-up days with survey data (n=8) or <6 follow-up days with samples with rRT-PCR results (n=14), co-primary case (n=56) or contacts in households with a co-primary case (n=46), and no remaining eligible household contacts after other exclusions (n=62 primary cases; Fig 1).

**Figure 1.**
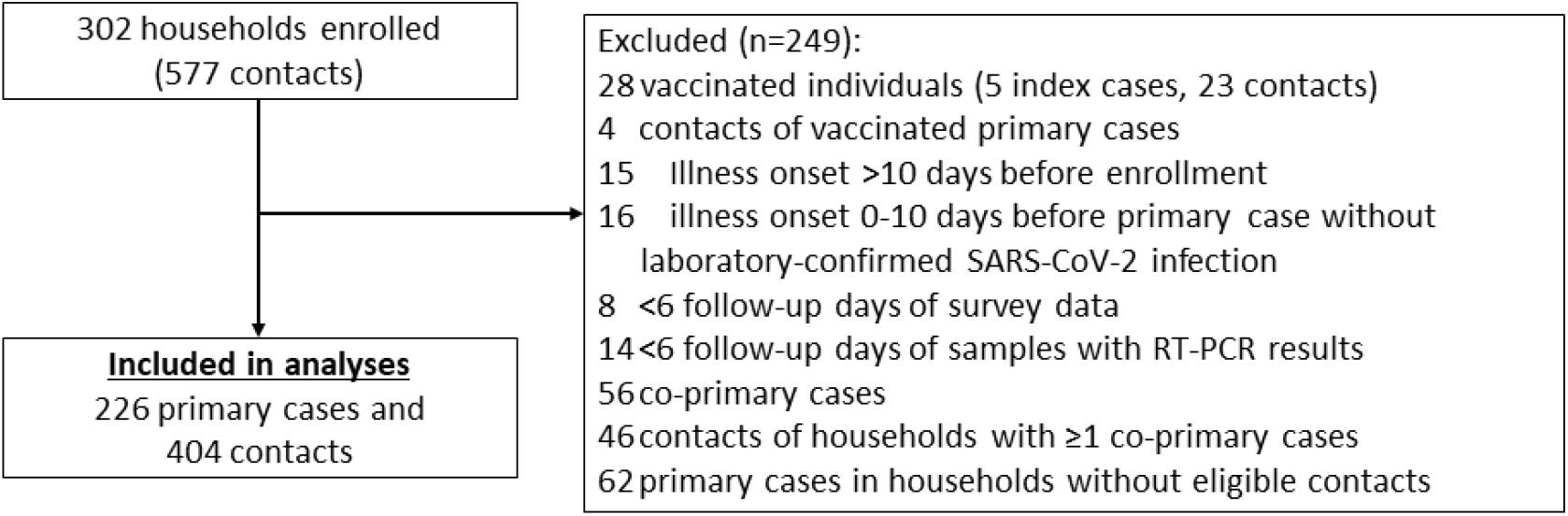
Households and individuals enrolled in a prospective study of SARS-CoV-2 household transmission — Tennessee and Wisconsin, April 2020–April 2021.

The majority of primary cases were non-Hispanic White young adults with a preponderance of females (Table 1). Median age was 37 years (range: 1-76 years). An underlying medical condition was reported by 81 (36%), of whom 29 (36%) had asthma. Among adults, 20% reported working in a healthcare setting and had regular face-to-face contact with sick people and 19% in customer service with regular face-to-face contact with people. Most primary cases (83%) lived in a single-family home with mean of 3.2 bedrooms and mean of 3.3 members. Among those who reported on interactions with other household members, interactions decreased from the day before illness onset to the day before enrollment; 73% reported physical contact with ≥1 other household member the day before illness onset versus 38% the day before enrollment (Table 1). Masking when interacting with other members was uncommon before illness onset (6%) and 26% reported mask use the day before enrollment.

**Table 1.** Characteristics of primary cases by age group at enrollment in a prospective study of SARS-CoV-2 household transmission — Tennessee and Wisconsin, April 2020–April 2021

|  | Age |  |  |  |  |  |  |
| --- | --- | --- | --- | --- | --- | --- | --- |
|  | 0-4 years | 5-11 years | 12-17 years | 18-49 years | 50-64 years | ≥65 years | All ages |
| **N (row %)** | 2 (0.9) | 4 (1.8) | 25 (11.1) | 141 (62.4) | 37 (16.4) | 17 (7.5) | 226 (100) |
| **Female** | 2 (100) | 0 (0) | 15 (60) | 81 (57) | 21 (57) | 9 (53) | 128 (56.6) |
| **Race/ethnicity<sup>1</sup>** |  |  |  |  |  |  |  |
| Non-Hispanic White | --- | --- | 23 (92) | 115 (82) | 35 (95) | 17 (100) | 194 (85.8) |
| Non-Hispanic other race | --- | --- | 0 | 9 (6) | 1 (3) | 0 | 11 (4.9) |
| Hispanic or Latino | --- | --- | 2 (8) | 16 (11) | 1 (3) | 0 | 20 (8.9) |
| **Smoker (aged ≥18 years)** | --- | --- | --- | 7 (5) | 2 (5) | 0 | 9 (5) |
| **Any underlying medical conditions<sup>3</sup>** | 1 (50) | 0 | 9 (36) | 46 (33) | 13 (35) | 12 (71) | 81 (35.8) |
| **Occupational, school, or childcare exposures** |  |  |  |  |  |  |  |
| Attended childcare or school outside home<sup>4</sup> (aged <18 years) | 1 (100) | 3 (100) | 14 (74) | --- | --- | --- | 18 (78) |
| Healthcare setting (aged ≥18 years)<sup>4,5</sup> | --- | --- | --- | 33 (24) | 6 (16) | 0 | 39 (20) |
| Customer service (aged ≥18 years)<sup>4,5</sup> | --- | --- | --- | 31 (22) | 4 (11) | 2 (12) | 37 (19) |
| Teacher (aged ≥18 years)<sup>4</sup> | --- | --- | --- | 4 (4) | 2 (7) | 1 (8) | 7 (5) |
| **Household characteristics** |  |  |  |  |  |  |  |
| Mean (SD) number of household members<sup>6</sup> | 4.0 (0) | 4.3 (1.7) | 4.4 (0.9) | 3.4 (1.4) | 2.7 (0.8) | 2.1 (0.2) | 3.3 (1.4) |
| Type of home |  |  |  |  |  |  |  |
| Single family home | 2 (100) | 3 (75) | 25 (100) | 110 (78) | 33 (89) | 14 (82) | 187 (82.7) |
| Duplex / townhome | 0 | 0 | 0 | 8 (6) | 0 | 0 | 8 (3.5) |
| Condo / apartment building | 0 | 1 (25) | 0 | 23 (16) | 4 (11) | 3 (18) | 31 (13.7) |
| Mean (SD) number of bedrooms<sup>6</sup> | 4.0 (0) | 3.3 (1.0) | 3.9 (0.7) | 3.0 (1.0) | 3.4 (1.2) | 2.9 (0.7) | 3.2 (1.0) |
| **Interactions with other household members<sup>4,7</sup>** |  |  |  |  |  |  |  |
| Maximum time spent in same room with ≥1 other member |  |  |  |  |  |  |  |
| Day before illness onset |  |  |  |  |  |  |  |
| >4 hours | 2 (100) | 2 (100) | 10 (43) | 56 (59) | 10 (40) | 6 (60) | 86 (55) |
| 1-4 hours | 0 | 0 | 7 (30) | 23 (24) | 11 (44) | 1 (10) | 42 (27) |
| <1 hour | 0 | 0 | 3 (13) | 16 (17) | 4 (16) | 3 (30) | 26 (17) |
| No time | 0 | 0 | 3 (13) | 0 | 0 | 0 | 3 (2) |
| Day before enrollment |  |  |  |  |  |  |  |
| >4 hours | 2 (100) | 1 (50) | 4 (17) | 35 (37) | 6 (23) | 2 (20) | 50 (32) |
| 1-4 hours | 0 | 0 | 7 (30) | 15 (16) | 6 (23) | 3 (30) | 31 (20) |
| <1 hour | 0 | 1 (50) | 8 (35) | 26 (27) | 7 (27) | 4 (40) | 46 (29) |
| No time | 0 | 0 | 4 (17) | 19 (20) | 7 (27) | 1 (10) | 31 (20) |
|  | 0-4 years | 5-11 years | 12-17 years | 18-49 years | 50-64 years | ≥65 years | All ages |
| **Had physical contact with ≥1 other members** |  |  |  |  |  |  |  |
| Day before illness onset | 2 (100) | 2 (100) | 15 (75) | 69 (73) | 19 (73) | 5 (56) | 112 (73) |
| Day before enrollment | 2 (100) | 1 (50) | 6 (32) | 37 (46) | 3 (14) | 2 (22) | 51 (38) |
| **Slept in the same room with ≥1 other members** |  |  |  |  |  |  |  |
| Day before illness onset | 0 | 1 (50) | 3 (15) | 43 (45) | 12 (46) | 5 (50) | 64 (41) |
| Day before enrollment | 0 | 0 | 1 (5) | 29 (36) | 5 (24) | 2 (22) | 37 (28) |
| **Frequency of masking when interacting with ≥1 other members** |  |  |  |  |  |  |  |
| Day before illness onset |  |  |  |  |  |  |  |
| Never | 1 (100) | --- | 7 (88) | 44 (98) | 15 (83) | 9 (100) | 76 (94) |
| Sometimes | 0 | --- | 1 (13) | 1 (2) | 3 (17) | 0 | 5 (6) |
| Always | 0 | --- | 0 | 0 | 0 | 0 | 0 |
| Day before enrollment |  |  |  |  |  |  |  |
| Never | 1 (100) | --- | 5 (63) | 33 (79) | 11 (65) | 7 (78) | 57 (74) |
| Sometimes | 0 | --- | 2 (25) | 3 (7) | 1 (6) | 0 | 6 (8) |
| Always | 0 | --- | 1 (13) | 6 (14) | 5 (29) | 2 (22) | 14 (18) |

Data are no. (% of column total with response) unless otherwise noted, numbers reflect rounding
Abbreviation: SD, standard deviation
<sup>1</sup>Unknown for 1 participant
<sup>2</sup>Data suppressed to protect privacy
<sup>3</sup>Underlying medical conditions included: asthma, cancer, cardiovascular or heart disease, diabetes, extreme obesity, high blood pressure or hypertension, immunocompromising condition, kidney disease, liver disease, other chronic lung disease, pregnancy, and prematurity
<sup>4</sup>Missing responses
<sup>5</sup>Healthcare setting were those who reported working in a healthcare setting and having regular face-to-face contact with sick people, customer service were those who reported working in customer service where they have regular face-to-face contact with people
<sup>6</sup>Data presented in this row are mean and standard deviation
<sup>7</sup>Restricted to primary cases who were also the index case in the household

Children aged 0-4 years and adults aged ≥65 years represented a minority (5% and 6%, respectively) of household contacts (Table 2). Other characteristics were similar to those reported among primary cases.

**Table 2.** Characteristics of household contacts by age group at enrollment in a prospective study of SARS-CoV-2 household transmission — Tennessee and Wisconsin, April 2020–April 2021

|  | Age |  |  |  |  |  |  |
| --- | --- | --- | --- | --- | --- | --- | --- |
|  | 0-4 years | 5-11 years | 12-17 years | 18-49 years | 50-64 years | ≥65 years | All ages |
| **N (row %)** | 21 (5.2) | 53 (13.1) | 67 (16.6) | 184 (45.5) | 53 (13.1) | 26 (6.4) | 404 (100) |
| **Female** | 9 (43) | 25 (47) | 28 (42) | 98 (53) | 28 (53) | 13 (50) | 201 (49.8) |
| **Race/Ethnicity** |  |  |  |  |  |  |  |
| **Non-Hispanic White** | 19 (90) | 40 (75) | 55 (82) | 148 (80) | 48 (91) | 24 (92) | 334 (82.7) |
| **Non-Hispanic other race** | 1 (5) | 2 (4) | 4 (6) | 19 (10) | 1 (2) | 2 (8) | 29 (7.2) |
| **Hispanic or Latino** | 1 (5) | 11 (21) | 8 (12) | 17 (9) | 4 (8) | 0 | 41 (10.2) |
| **Smoker (aged ≥18 years)** | --- | --- | --- | 12 (7) | 3 (6) | 0 | 15 (6) |
| **Any underlying medical conditions<sup>1</sup>** | 0 | 6 (11) | 15 (22) | 53 (29) | 17 (32) | 16 (62) | 107 (26.5) |
| **Occupational, school, or childcare exposures** |  |  |  |  |  |  |  |
| **Attended childcare or school outside home<sup>2</sup> (aged <18 years)** | 5 (56) | 21 (66) | 29 (71) | --- | --- | --- | 55 (67) |
| **Healthcare setting (aged ≥18 years)<sup>2,3</sup>** | --- | --- | --- | 17 (9) | 5 (9) | 1 (4) | 23 (8.8) |
| **Customer service (aged ≥18 years)<sup>2,3</sup>** | --- | --- | --- | 36 (20) | 10 (19) | 2 (8) | 48 (18.5) |
| **Teacher (aged ≥18 years)<sup>2</sup>** | --- | --- | --- | 8 (6) | 2 (5) | 1 (5) | 11 (5.5) |

Data are no. (% of column total with response) unless otherwise noted, numbers reflect rounding
<sup>1</sup>Underlying medical conditions included: asthma, cancer, cardiovascular or heart disease, diabetes, extreme obesity, high blood pressure or hypertension, immunocompromising condition, kidney disease, liver disease, other chronic lung disease, pregnancy, and prematurity
<sup>2</sup>Missing responses
<sup>3</sup>Healthcare setting were those who reported working in a healthcare setting and having regular face-to-face contact with sick people, customer service were those who reported working in customer service where they have regular face-to-face contact with people

### Transmission from primary cases and infection among contacts

The 226 primary cases were followed by 198 (49%) SARS-CoV-2 infections among 404 household contacts. At least one contact was infected in 58% (130 of 226) of households. Estimated SIR ranged from 26% among contacts of primary cases aged 12-17 years to 76% among contacts of primary cases aged ≥65 years (Fig 2). Compared to when the primary case was aged 18-49 years, SIR in household contacts was significantly lower when the primary case was aged 12-17 years (RR 0.42; 95% CI 0.19-0.91), and not significantly different for all other primary case age groups. There were no significant differences in estimated SIR by age of the contacts (Fig 2). SIR ranged from 36% among contacts aged ≥65 years to 53% among contacts aged 5-11 years.

**Figure 2.**
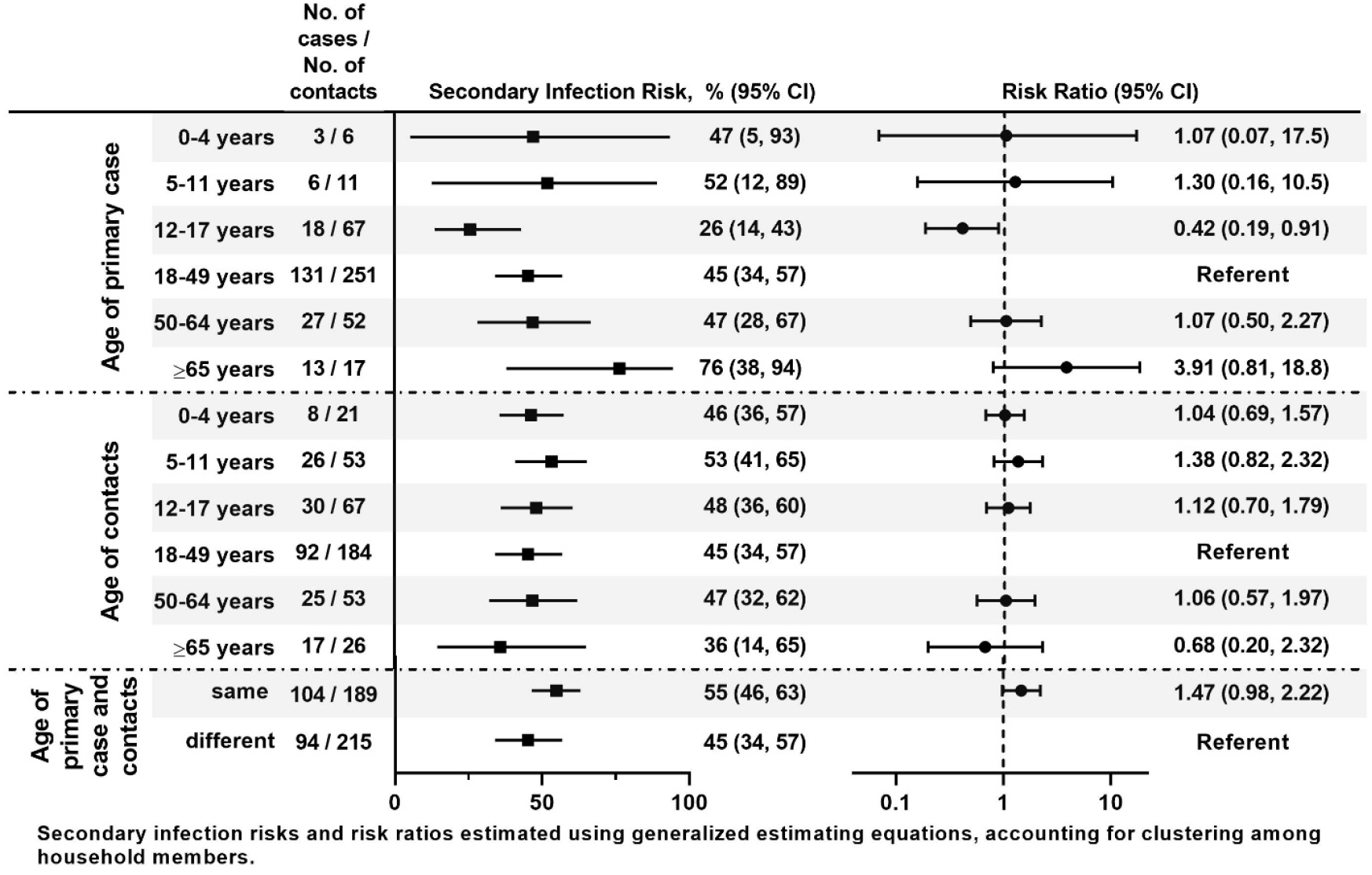
Estimated transmission risk from the primary case and infection risk among household contacts by age — Prospective study of SARS-CoV-2 household transmission, Tennessee and Wisconsin, April 2020–April 2021.

Overall, estimated SIR was higher when primary case-contact pairs were in the same versus different age groups (55% versus 45%, RR 1.47; 95% CI 0.98-2.22; Fig 2). SIR was highest among primary case-contacts pairs aged ≥65 years (76%) and 5-11 years (69%). Within each primary case age group, SIR was generally lowest among contacts aged ≥65 years (Fig 3).

**Figure 3.**
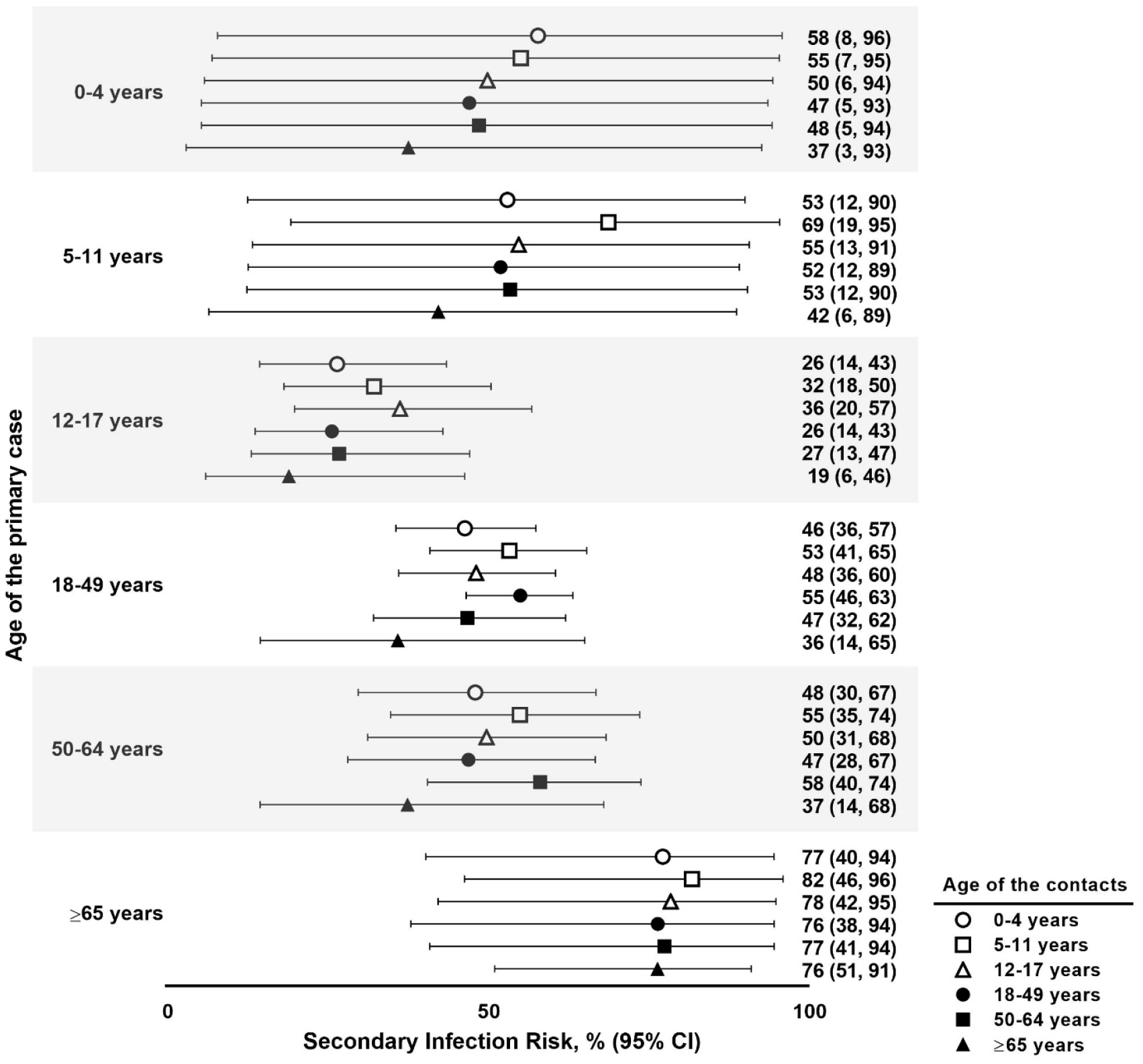
Estimated secondary infection risk by age of the primary case and age of the household contacts — Prospective study of SARS-CoV-2 household transmission, Tennessee and Wisconsin, April 2020–April 2021.

However, CIs were wide for all primary case-contact age group combinations, particularly those aged <12 years.

### Symptoms among contacts with SARS-CoV-2 infection

Among 186 contacts with SARS-CoV-2 infection with symptom onset or first positive rRT-PCR result after enrollment, 96% completed symptom surveys on ≥13 days of follow-up (Table 3). Most (81%) reported ≥1 symptoms and 19% reported no symptoms (asymptomatic) (Fig 4). There were no differences in asymptomatic infections by age group (ranging from 12% among those aged 50-64 years to 27% among those aged 5-11 years). Among the 150 symptomatic infections, 46% were rRT-PCR positive (from nasal or saliva sample) before symptom onset. Median days from first positive viral detection to symptom onset was 2 (interquartile range [IQR] 1-3) and did not differ by age group (*P*=0.90). Median number of days with a positive rRT-PCR result (from nasal samples) during follow-up was 8 (IQR 4-11); however, 34% continued to be positive on the last day of follow-up.

**Table 3.** Characteristics of contacts with SARS-CoV-2 infection included in symptom analysis by age group — Prospective study of SARS-CoV-2 household transmission, Tennessee and Wisconsin, April 2020–April 2021

|  | 0-4 years | 5-11 years | 12-17 years | 18-49 years | 50-64 years | ≥65 years | All ages |
| --- | --- | --- | --- | --- | --- | --- | --- |
| **N (row %)** | 8 (4) | 26 (14) | 30 (16) | 81 (44) | 25 (13) | 16 (9) | 186 (100) |
| **Female, n (column %)** | 3 (38) | 11 (42) | 15 (50) | 36 (44) | 11 (44) | 10 (63) | 86 (46) |
| **Race/ethnicity, n (column %)** |  |  |  |  |  |  |  |
| Non-Hispanic White | 8 (100) | 20 (77) | 24 (80) | 63 (78) | 23 (92) | 16 (100) | 154 (83) |
| Non-Hispanic other race | 0 | 0 | 2 (7) | 8 (10) | 0 | 0 | 10 (5) |
| Hispanic or Latino | 0 | 6 (23) | 4 (13) | 10 (12) | 2 (8) | 0 | 22 (12) |
| **Smoker (aged ≥18 years), n (column %)** | --- | --- | --- | 2 (2) | 1 (4) | 0 | 3 (2) |
| **Any underlying medical conditions<sup>1</sup>, n (column %)** | 0 | 2 (8) | 1 (3) | 20 (25) | 9 (36) | 10 (63) | 42 (23) |
| **Occupational, school, or childcare exposures, n (column %)** |  |  |  |  |  |  |  |
| Attended childcare or school outside home<sup>2</sup> (aged <18 years) | 2 (50) | 15 (65) | 11 (55) | --- | --- | --- | 28 (60) |
| **Healthcare setting (aged ≥18 years)<sup>2,3</sup>** | --- | --- | --- | 6 (8) | 3 (12) | 1 (6) | 10 (8) |
| **Customer service (aged ≥18 years)<sup>2,3</sup>** | --- | --- | --- | 14 (18) | 4 (16) | 2 (13) | 20 (17) |
| **Teacher (aged ≥18 years)<sup>2</sup>** | --- | --- | --- | 4 (6) | 1 (5) | 1 (8) | 6 (6) |
| **Completed ≥13 (of 14) symptom surveys, n (column %)** | 8 (100) | 26 (100) | 29 (97) | 80 (99) | 24 (96) | 12 (75) | 179 (96) |
| **Took medication for fever or pain at least 1 day, n (column %)** | 6 (75) | 9 (35) | 11 (37) | 41 (51) | 18 (72) | 9 (56) | 94 (51) |
| **Sought medical care during follow-up, n (column %)<sup>2</sup>** | 0 | 0 | 2 (7) | 5 (6) | 0 | 2 (13) | 9 (5) |
| **Timing of positive result relative to symptom onset** |  |  |  |  |  |  |  |
| Positive rRT-PCR result before symptom onset, n (column %)<sup>4,6</sup> | 3 (43) | 11 (58) | 10 (42) | 30 (46) | 11 (50) | 4 (31) | 69 (46) |
| Median (IQR) days from first positive to symptom onset<sup>4,5,6</sup> | 2 (1, 3) | 3 (1, 3) | 2 (1, 3) | 1.5 (1, 3) | 2 (1, 4) | 2 (1, 5) | 2 (1, 3) |
| **Median (IQR) days positive during follow-up period<sup>7</sup>** | 9 (6.5, 11) | 8 (3.5, 10.5) | 8 (6, 11) | 8 (4, 11) | 12 (7, 13) | 8 (1, 12) | 8 (4, 11) |
| **Median (IQR) days from primary case onset** |  |  |  |  |  |  |  |
| To first positive | 7 (6.5, 9) | 6 (5, 9) | 6 (5, 8) | 6 (4, 8) | 5 (5, 7) | 5 (4, 5.5) | 6 (4, 8) |
| To first symptom<sup>4</sup> | 8 (6, 8) | 6 (5, 10) | 6 (4.5, 9) | 6 (4, 7) | 6 (5, 7) | 5 (4, 6) | 6 (5, 8) |

Abbreviation: IQR, interquartile range; rRT-PCR, real-time reverse transcription polymerase chain reaction
<sup>1</sup>Underlying medical conditions included: asthma, cancer, cardiovascular or heart disease, diabetes, extreme obesity, high blood pressure or hypertension, immunocompromising condition, kidney disease, liver disease, other chronic lung disease, pregnancy, and prematurity
<sup>2</sup>Missing responses
<sup>3</sup>Healthcare setting were those who reported working in a healthcare setting and having regular face-to-face contact with sick people, customer service were those who reported working in customer service where they have regular face-to-face contact with people
<sup>4</sup>Among contacts who reported any symptoms. Symptoms assessed included: constitutional symptoms (chills, fatigue or feeling run down, fever or feverishness, muscle or body aches), upper respiratory symptoms (nasal congestion, runny nose, sore throat), lower respiratory symptoms (chest tightness or pain, cough,
trouble breathing or shortness of breath, wheezing), neurologic symptoms (headache, loss of taste or smell), and gastrointestinal symptoms (abdominal pain, diarrhea, vomiting). Gastrointestinal symptoms were assessed in Wisconsin only
<sup>5</sup>Among contacts with a positive rRT-PCR result before symptom onset
<sup>6</sup>Impacted by left-censoring, participants may have been positive prior to their symptom onset and positive prior to the start of follow-up
<sup>7</sup>Restricted to nasal samples only; impacted by censoring (left and right)

**Figure 4.**
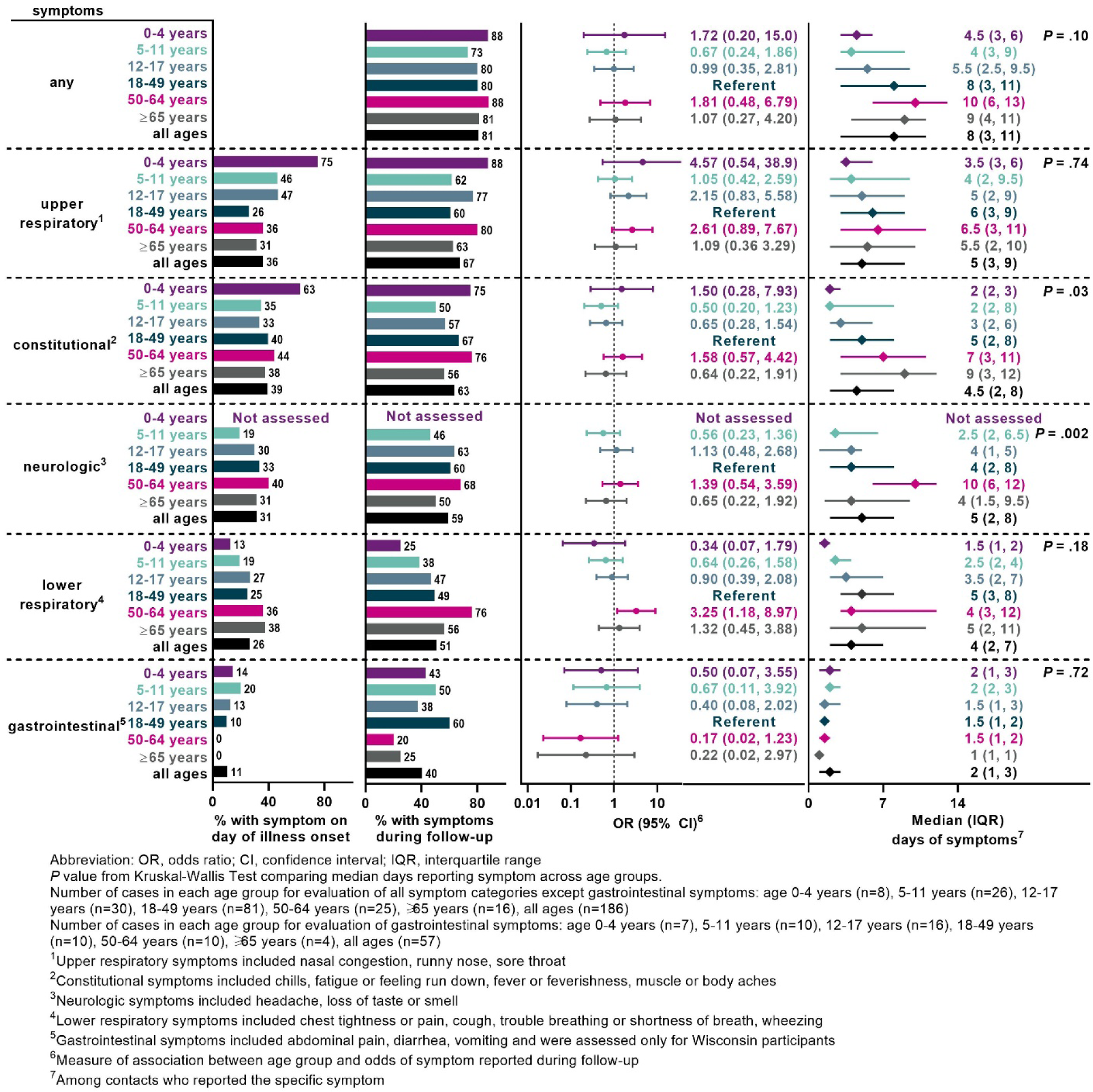
Reported symptoms, timing, and duration of symptoms by age group among persons with SARS-CoV-2 infection in a prospective study of SARS-CoV-2 household transmission — Tennessee and Wisconsin, April 2020–April 2021.

Among infected children, the most commonly reported symptoms were upper respiratory symptoms (88% aged 0-4 years, 62% aged 5-11 years, and 77% aged 12-17 years; Fig 4). Lower respiratory symptoms were reported by 68% (28 of 41) of adults aged ≥50 years (76% aged 50-64 years, 56% aged ≥65 years). Gastrointestinal symptoms were less common than other symptoms (40%) and not typically reported on the day of illness onset, but was commonly reported in children aged 5-11 years (50%) and adults aged 18-49 years (60%). Frequency and duration of individual symptoms are shown in Supplemental Fig.

Median duration of any symptom was 8 days (IQR 3-11) and did not differ by age group; 34% reported symptoms on the last day of follow-up and thus their symptom duration was foreshortened. However, median duration of constitutional symptoms was significantly different and increased with age, from 2 days in younger children (age <12 years) to 9 days in adults aged ≥65 years (*P*=0.03). Median duration of neurologic symptoms was ≤4 days for all age groups except adults aged 50-64 years, where median of 10 days was observed (*P*=0.002).

## DISCUSSION

This prospective study examined the association of age with household transmission of SARS-CoV-2, addressing both age of the primary case (transmission risk) and ages of household contacts (susceptibility). With frequent and systematic testing among household contacts, we found much higher infection risks among household contacts than prior studies, similar rates of infection among child and adult household contacts, and some nuanced differences in transmission from specific age groups. However, transmission to household contacts was observed from primary cases in all age groups.

There is considerable heterogeneity in the literature on transmission from and susceptibility of children to SARS-CoV-2. Our finding of similar infection risk across age groups is consistent with larger investigations of reported cases from Spain,^18^ China,^6^ and Brunei^19^ and other household transmission studies conducted in the US.^20^ However, several studies have reported that children had reduced susceptibility to SARS-CoV-2 infection^21-24^ and one found higher risk in children compared with young adults.^25^ Difference in methods for ascertaining and detecting SARS-CoV-2 infection among household contacts may account for some of the differences between studies. Infrequent or delayed testing, or limited testing of asymptomatic/mildly-symptomatic individuals could contribute to underestimation of SARS-CoV-2 infections and may differentially impact infection risk measurements among children, thus biasing the association between age and susceptibility and transmission.

Additionally, some prior studies did not disaggregate age into finer age groups, analyzing all children aged <18 years together,^5,9,20,24,26-29^ which may obscure biological or behavioral characteristics that vary by age. We categorized our study participants into six age groups, roughly aligned with US groupings in school (preschool, primary school, and secondary school) and adulthood periods (young, middle-aged, and older adults). In doing so, we found that transmission risk was highest from primary cases aged ≥65 years and lowest from primary cases aged 12-17 years. Studies from Ontario and Denmark also found transmission risk in children was highest among the youngest and lowest in adolescents.^30,31^ Age-specific differences in behavior likely contributed to lower transmission from adolescents relative to younger, less autonomous, children.

Other studies have also identified somewhat increased transmission risk among contacts of similar age, and this may have implications beyond the household.^31,32^ Transmission patterns may be influenced by assortative mixing, where similar aged people interact more with each other than with different aged people.^33,34^ These findings may inform planning for school-based countermeasures to reduce transmission risk within and between classrooms. However, further investigation is needed to better understand how behaviors and interactions differ by age, during periods of illness, and whether those differences are associated with risk of transmission in both the household and school settings.

In this study and others,^1^ young children aged <12 years were rarely identified as the primary case of SARS-CoV-2 infection in households. However, when a young child was the primary case, we saw that they transmitted infection to more than 37% of their household contacts, including adults, and their probability of transmission in the household setting was not significantly different than transmission from young adults. Similar rates of transmissibility between children and adults were also found in studies conducted in China and South Korea,^21,35^ though small sample size in young children also limited those studies.

The frequency and duration of symptoms were similar across age groups among those who were infected in our study. Clinical data from non-hospitalized infected children are limited, with few studies directly comparing symptoms of children and adults.^5,20,36,37^ There were no significant differences in the frequency of asymptomatic infections between children and adults. The overall percentage of infected participants reporting no symptoms is generally consistent with the 20% estimated in a recent review and meta-analysis.^38^ Additional studies to better understand the frequency of asymptomatic infections by age and the role of asymptomatic infection and age in onward transmission in household and community settings are needed to inform public health recommendations.

This study has several limitations. First, delayed identification of index cases prevented complete capture of transmission events. Participants may have become infected, but remain asymptomatic, between the time the index case was tested and study enrollment. Thus, duration of positivity and symptoms captured during the enrollment period may be subject to both right and left censoring and may be an underestimate. Second, we assumed secondary infections among contacts resulted from household exposure rather than community transmission. Ongoing exposure from the community may lead to an overestimation of transmission in household settings, especially among age groups more likely to be exposed outside the household. Although patients were instructed to isolate at home or quarantine while waiting for results,^39^ compliance may have declined as the pandemic progressed. Third, we did not account for reported interactions between the primary case and contacts before and during illness in the primary case. While physical contact between the primary case and household contacts was common, mask use by the primary case was not common before or after illness onset. Differences in age-related interactions may explain or help clarify associations between age and transmission events. Fourth, the study population was largely non-Hispanic White, and findings may not be generalizable to other racial and ethnic groups. Finally, small sample size limited the precision of estimates of SIR and our statistical power to detect true differences in transmission risk and symptom profiles by age. Despite these limitations, the case-ascertained household study described provides much needed evidence regarding susceptibility to SARS-CoV-2 infection, as exposure within the household is well-defined and prolonged. Additionally, daily follow-up allowed us to assess symptoms throughout the course of illness.

In conclusion, we observed that both children and adults of all ages can transmit and are susceptible to SARS-CoV-2 infection. There were no significant differences in susceptibility to SARS-CoV-2 by age group, from preschool-aged children through older adults. Further research is needed to understand age-related interactions and behaviors in households as it relates to the probability of transmission by age.

## Data Availability

Deidentified individual participant data (including data dictionaries) will be made available upon request after publication to researchers who provide a methodologically sound proposal for use in achieving the goals of the approved proposal. Proposals should be submitted to.

## Acknowledgements

We thank the following for their contributions to the study: Lynn Ivacic, Hannah Berger, Vicki Moon, Keegan Brighton, Gina Burbey, Deanna Cole, Leila Deering, Eric DeJarlais, Heather Dirkx, Sherri Guzinski, Joshua Hebert, Linda Heeren, Erin Higdon, Jacob Johnston, Chris Kadolph, Taylor Kent, Burney Kieke, Tamara Kronenwetter Koepel, Sarah Kohn, Diane Kohnhorst, Erik Kronholm, Stacey Kyle, Jim Linneman, Carrie Marcis, Karen McGreevey, Sudha Medabalimi, Nidhi Mehta, Nan Pan, Cory Pike, Rebecca Pilsner, DeeAnn Polacek, Martha Presson, Carla Rottscheit, Jacklyn Salzwedel, Kristin Seyfert, Tapan Sharma, Alyssa Spoerl, Sandy Strey, Krishna Chaitanya Upadhyay, Gail Weinand, and Benjamin Zimmerman at Marshfield Clinic Research Institute; Judy King, Dayna Wyatt, Robert Lyons, Carleigh Frazier, Emily Jookar, Karen Malone, Olivia Doak, Sarah Davis, Jorge Celedonio, Marcia Blair, Rendie McHenry, Claudia Guevara, Jennifer Luther, Laura Short, and Ahra Kim at Vanderbilt University Medical Center.

## Abbreviations

CDC: Centers for Disease Control and Prevention
CI: Confidence interval
GEE: General estimating equations
IQR: Interquartile range
MCHS: Marshfield Clinic Health System
OR: Odds ratio
RR: Risk ratio
rRT-PCR: Real-time reverse transcription polymerase chain reaction
SD: Standard deviation
SIR: Secondary infection risk
US: United States
VUMC: Vanderbilt University Medical Center

## References

1. Viner RM, Mytton OT, Bonell C, et al. Susceptibility to SARS-CoV-2 infection among children and adolescents compared with adults: a systematic review and meta-analysis. JAMA Pediatr 2021;175:143–56.

2. Goldstein E, Lipsitch M, Cevik M. On the effect of age on the transmission of SARS-CoV-2 in households, schools, and the community. J Infect Dis 2021;223:362–9.

3. Spielberger BD, Goerne T, Geweniger A, Henneke P, Elling R. Intra-household and close-contact SARS-CoV-2 transmission among children - a systematic review. Front Pediatr 2021;9:613292.

4. Madewell ZJ, Yang Y, Longini IM, Jr., Halloran ME, Dean NE. Household transmission of SARS-CoV-2: a systematic review and meta-analysis. JAMA Netw Open 2020;3:e2031756.

5. Yousaf AR, Duca LM, Chu V, et al. A prospective cohort study in non-hospitalized household contacts with SARS-CoV-2 infection: symptom profiles and symptom change over time. Clin Infect Dis 2020.

6. Bi Q, Wu Y, Mei S, et al. Epidemiology and transmission of COVID-19 in 391 cases and 1286 of their close contacts in Shenzhen, China: a retrospective cohort study. Lancet Infect Dis 2020;20:911–9.

7. Accorsi EK, Qiu X, Rumpler E, et al. How to detect and reduce potential sources of biases in studies of SARS-CoV-2 and COVID-19. Eur J Epidemiol 2021;36:179–96.

8. Mehta NS, Mytton OT, Mullins EWS, et al. SARS-CoV-2 (COVID-19): what do we know about children? A systematic review. Clin Infect Dis 2020;71:2469–79.

9. Liguoro I, Pilotto C, Bonanni M, et al. SARS-COV-2 infection in children and newborns: a systematic review. Eur J Pediatr 2020;179:1029–46.

10. Havers FP, Whitaker M, Self JL, et al. Hospitalization of adolescents aged 12-17 years with laboratory-confirmed COVID-19 - COVID-NET, 14 States, March 1, 2020-April 24, 2021. MMWR Morb Mortal Wkly Rep 2021;70:851–7.

11. Estimated disease burden of COVID-19. 2021. (Accessed June 28, 2021, at https://www.cdc.gov/coronavirus/2019-ncov/cases-updates/burden.html.)

12. Grijalva CG, Rolfes MA, Zhu Y, et al. Transmission of SARS-COV-2 infections in households - Tennessee and Wisconsin, April-September 2020. MMWR Morb Mortal Wkly Rep 2020;69:1631–4.

13. Rolfes MA, Grijalva CG, Zhu Y, et al. Implications of shortened quarantine among household contacts of index patients with confirmed SARS-CoV-2 infection - Tennessee and Wisconsin, April-September 2020. MMWR Morb Mortal Wkly Rep 2021;69:1633–7.

14. COVID-19: get tested. 2021. (Accessed 7/19, 2021, at https://www.dhs.wisconsin.gov/covid-19/testing.htm.)

15. Centers for Disease Control and Prevention. CDC 2019-novel coronavirus (2019-nCoV) real-time RT-PCR diagnostic panel. 2020.

16. Quidel Corporation. Lyra® SARS-CoV-2 assay instructions for use. 2020.

17. ThermoFisher Scientific. TaqPath™ COVID-19 combo kit and TaqPath™ COVID-19 combo kit advanced* instructions for use. 2020.

18. Arnedo-Pena A, Sabater-Vidal S, Meseguer-Ferrer N, et al. COVID-19 secondary attack rate and risk factors in household contacts in Castellon (Spain): preliminary report. Rev Enf Emerg 2020;19:64–70.

19. Chaw L, Koh WC, Jamaludin SA, Naing L, Alikhan MF, Wong J. Analysis of SARS-CoV-2 transmission in different settings, Brunei. Emerg Infect Dis 2020;26:2598–606.

20. Laws RL, Chancey RJ, Rabold EM, et al. Symptoms and transmission of SARS-CoV-2 among children - Utah and Wisconsin, March-May 2020. Pediatrics 2021;147.

21. Hu S, Wang W, Wang Y, et al. Infectivity, susceptibility, and risk factors associated with SARS-CoV-2 transmission under intensive contact tracing in Hunan, China. Nat Commun 2021;12:1533.

22. Li F, Li YY, Liu MJ, et al. Household transmission of SARS-CoV-2 and risk factors for susceptibility and infectivity in Wuhan: a retrospective observational study. Lancet Infect Dis 2021;21:617–28.

23. Dattner I, Goldberg Y, Katriel G, et al. The role of children in the spread of COVID-19: using household data from Bnei Brak, Israel, to estimate the relative susceptibility and infectivity of children. PLoS Comput Biol 2021;17:e1008559.

24. Jing QL, Liu MJ, Zhang ZB, et al. Household secondary attack rate of COVID-19 and associated determinants in Guangzhou, China: a retrospective cohort study. Lancet Infect Dis 2020;20:1141–50.

25. Liu T, Liang W, Zhong H, et al. Risk factors associated with COVID-19 infection: a retrospective cohort study based on contacts tracing. Emerg Microbes Infect 2020;9:1546–53.

26. Cheng HY, Jian SW, Liu DP, et al. Contact tracing assessment of COVID-19 transmission dynamics in Taiwan and risk at different exposure periods before and after symptom onset. JAMA Intern Med 2020;180:1156–63.

27. Lewis NM, Chu VT, Ye D, et al. Household Transmission of SARS-CoV-2 in the United States. Clin Infect Dis 2020.

28. Luo L, Liu D, Liao X, et al. Contact settings and risk for transmission in 3410 close contacts of patients with COVID-19 in Guangzhou, China : a prospective cohort study. Ann Intern Med 2020;173:879–87.

29. Wang Y, Tian H, Zhang L, et al. Reduction of secondary transmission of SARS-CoV-2 in households by face mask use, disinfection and social distancing: a cohort study in Beijing, China. BMJ Glob Health 2020;5.

30. Paul LA, Daneman N, Schwartz KL, et al. Association of Age and Pediatric Household Transmission of SARS-CoV-2 Infection. JAMA Pediatr 2021.

31. Lyngse FP, Mølbak K, Træholt Frank K, Nielsen C, Skov RL, Kirkeby CT. Association between SARS-CoV-2 transmission risk, viral load, and age: a nationwide study in Danish households. medRxiv 2021.

32. Laxminarayan R, Wahl B, Dudala SR, et al. Epidemiology and transmission dynamics of COVID-19 in two Indian states. Science 2020;370:691–7.

33. Mossong J, Hens N, Jit M, et al. Social contacts and mixing patterns relevant to the spread of infectious diseases. PLoS Med 2008;5:e74.

34. Hoang T, Coletti P, Melegaro A, et al. A systematic review of social contact surveys to inform transmission models of close-contact infections. Epidemiology 2019;30:723–36.

35. Park YJ, Choe YJ, Park O, et al. Contact tracing during coronavirus disease outbreak, South Korea, 2020. Emerg Infect Dis 2020;26:2465–8.

36. CDC COVID-19 Response Team. Coronavirus disease 2019 in children - United States, February 12-April 2, 2020. MMWR Morb Mortal Wkly Rep 2020;69:422–6.

37. Chung E, Chow EJ, Wilcox NC, et al. Comparison of symptoms and RNA levels in children and adults with SARS-CoV-2 infection in the community setting. JAMA Pediatr 2021.

38. Buitrago-Garcia D, Egli-Gany D, Counotte MJ, et al. Occurrence and transmission potential of asymptomatic and presymptomatic SARS-CoV-2 infections: A living systematic review and meta-analysis. PLoS Med 2020;17:e1003346.

39. Coronavirus disease 2019 (COVID-19) patient testing. 2021. (Accessed 7/19, 2021, at https://www.marshfieldclinic.org/patient-resources/patient-testing.)

