## Supplemental Fig for "Household Transmission and Clinical Features of SARS-CoV-2 Infections by Age in 2 US Communities"

**Supplemental Figure.** Reported symptoms, timing, and duration of specific symptoms by age group among persons with SARS-CoV-2 infection in a prospective study of SARS-CoV-2 household transmission — Tennessee and Wisconsin, April 2020–April 2021.

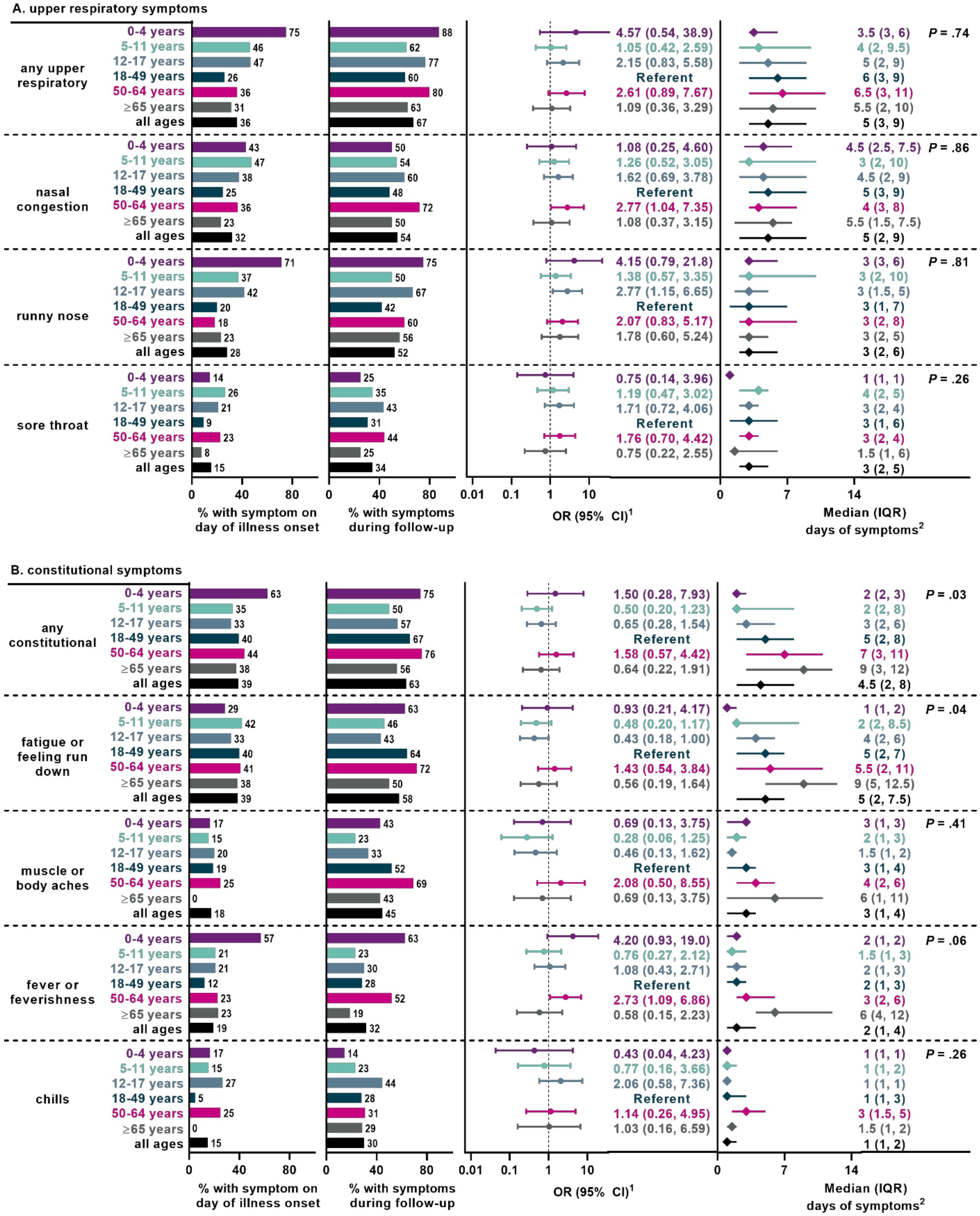

C. neurologic symptoms

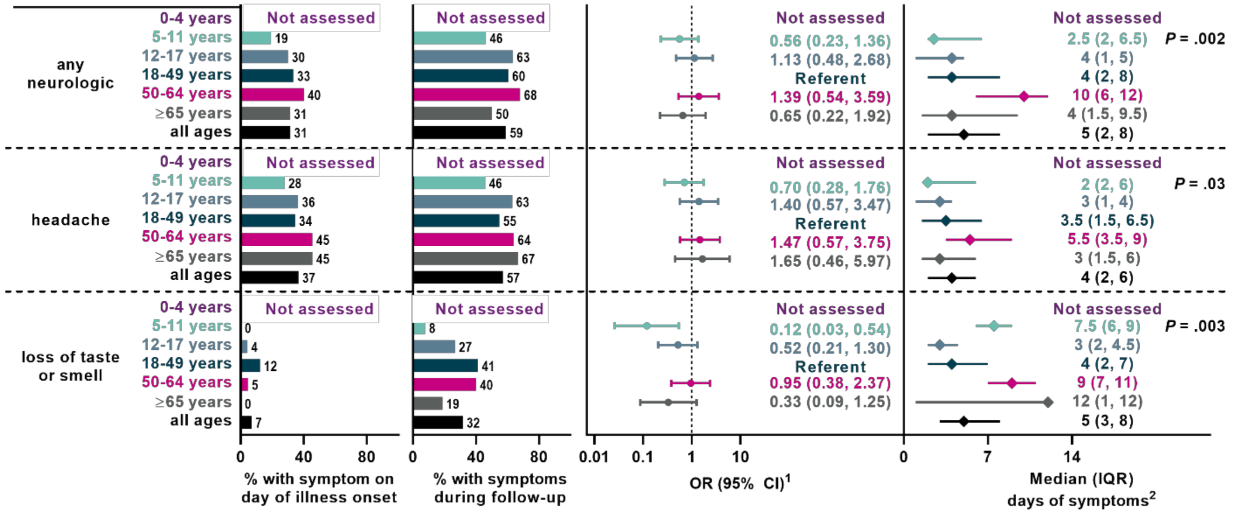


D. lower respiratory symptoms

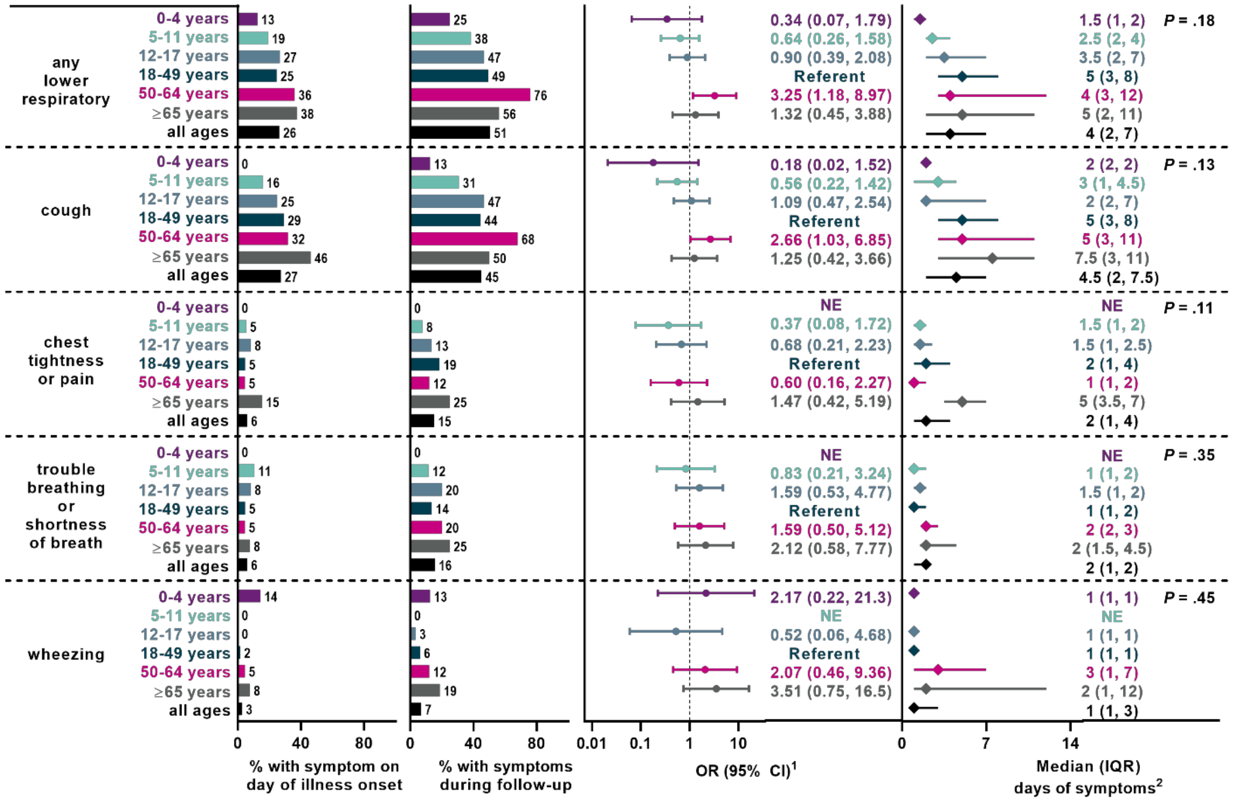

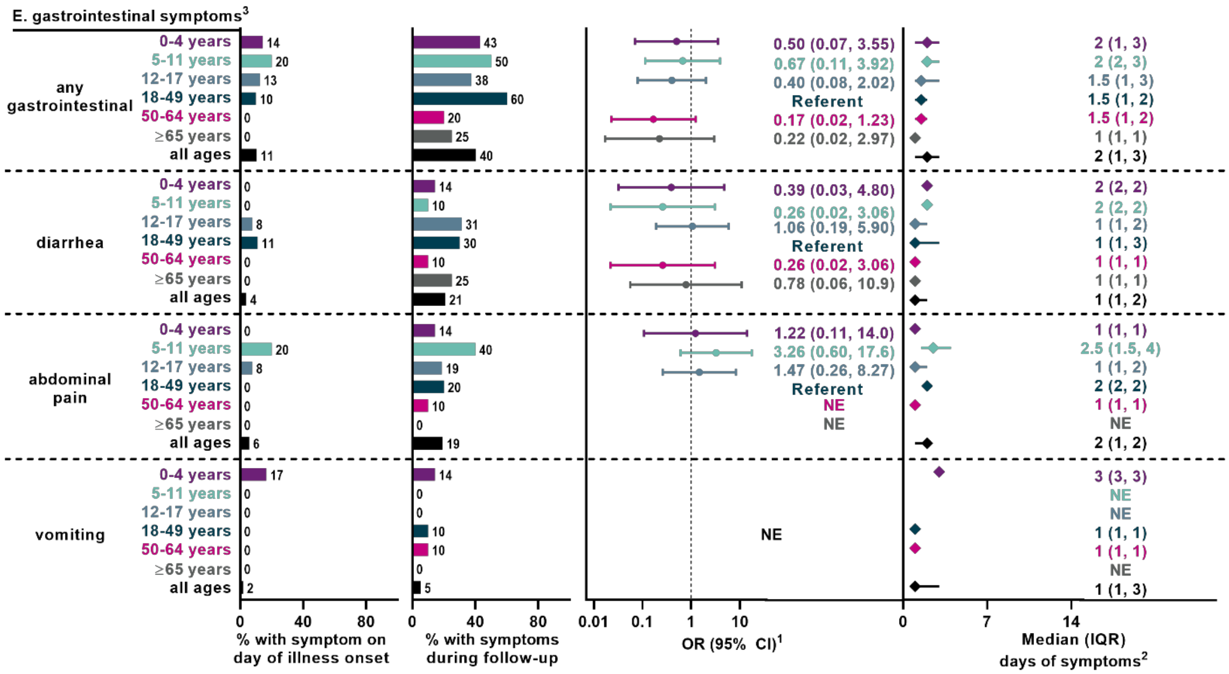


Abbreviation: OR, odds ratio; CI, confidence interval; IQR, interquartile range; NE, not estimated

*P* value from Kruskal-Wallis Test comparing median days with symptoms across age groups.

[1]Measure of association between age group and odds of symptom reported during follow-up

[2]Among contacts who reported the specific symptoms

[3]Gastrointestinal symptoms were assessed only for Wisconsin participants
